# Challenges in the medical oxygen ecosystem of Peru: a political economy analysis

**DOI:** 10.1101/2025.01.02.25319915

**Authors:** Patricia J. Garcia, Freddy Eric Kitutu, Jehnette M. Guzman, Lizzette Najarro, Freddie Ssengooba, Carina King

## Abstract

**Background:** Medical oxygen is essential in the management of several human disease conditions including acute respiratory conditions across the life course, and yet access remains unequal in many low- and middle-income countries, including Peru. This study explores Peru’s challenges in ensuring reliable oxygen supplies, with a focus on those laid bare or exacerbated by the COVID-19 pandemic, to inform strategies for improving medical oxygen access.

**Methodology and Principal Findings:** Using a political economy analysis, we conducted 13 key informant interviews with stakeholders involved in oxygen policy, supply, and health care, supported by reviews of 117 academic and grey literature sources, including policy documents. Before the COVID-19 pandemic, Peru’s requirement for medical oxygen to be >99% pure restricted competition, consolidating control of a few large liquid oxygen suppliers on the oxygen market and blocking smaller, affordable providers due to high compliance costs. Although pre-pandemic oxygen supplies were reportedly adequate, the pandemic exposed severe limitations including market constraints, slow government response, and lack of data management, resulting in an acute oxygen crisis. Civil society and private organizations stepped in, donating medical oxygen generator plants, but many of these are now unused due to insufficient planning for maintenance and operation.

**Conclusions and Significance:** This study underscores the urgent need for a National Oxygen System in Peru to oversee supply, distribution, and maintenance, and strengthen resilience for future health emergencies. Solutions include reducing reliance on a small number of external suppliers, infrastructure investments, dedicated funding for maintenance, and training for personnel to ensure continuous oxygen access nationwide. This research highlights systemic vulnerabilities in Peru’s health system and calls for coordinated policies to ensure equitable oxygen access and preparedness for future crises.

## Introduction

Medical oxygen is a critical yet often overlooked medicine, essential for treating a range of conditions, including respiratory illnesses and critical conditions. It has been in clinical use since the late 1800s (1). In 2002, the World Health Organization (WHO) included oxygen in the essential medicine list (EML) – but it was limited to anesthetic use, and only in 2017 was the indication to be used for hypoxemia added (2).

A large and diverse patient population relies on oxygen therapy, spanning from vulnerable newborns in respiratory distress to children with lower respiratory infections like pneumonia or bronchiolitis, sepsis, and congenital or acquired heart disease(3). Adults also face critical oxygen needs due to infectious causes of hypoxemia (such as tuberculosis, malaria, pneumonia, and HIV/AIDS), chronic respiratory illnesses (like COPD, asthma, pulmonary hypertension, and interstitial lung disease), and other hypoxemic conditions including ARDS, trauma, heart failure, post-surgical recovery, resuscitated sepsis, and cancer(4). The onset of the COVID-19 pandemic led to a substantial increase in the demand for oxygen due to the respiratory complications associated with the virus and highlighted the severe gaps and inequities in oxygen supply across health systems globally (5,6). The acute oxygen shortages experienced during the COVID-19 pandemic emerged as a critical disparity, underscoring the lack of preparedness and security in essential healthcare resources worldwide (7–9).

Peru is a Latin American (LA) middle-income country of roughly 34 million inhabitants of which around 2.9 million are under five years of age (10). In the two decades preceding the COVID-19 pandemic, Peru experienced steady economic growth and expanded healthcare access, yet significant socio-economic, regional, and ethnic disparities in healthcare access persisted (11–13). Life expectancy saw a significant increase by 2019, rising to 81.8 years for women and 78.7 years for men, compared to 71.6 and 67 years in 1990. This improvement is largely attributed to advancements in maternal and child healthcare and a substantial decline in mortality from infectious diseases (14).

The surge in COVID-19 cases in the country stressed the national healthcare system, revealing a lack of preparedness in oxygen availability and distribution affecting the management of cases and mortality. According to data from the Global Burden of Disease study, Peru experienced the highest excess mortality rate due to COVID-19 in 2020, with 413.4 excess deaths per 100,000 population (range: 410.3–416.1). As a result, life expectancy in 2021 regressed closer to 1990 levels, falling to 74.9 years for women and 68.8 years for men (15).

This paper examines the structural and policy-related obstacles Peru faced in ensuring adequate access to medical oxygen for those who need it, providing key insights for the global political economy of oxygen presented in the Lancet Global Health Commission on Medical Oxygen Security (16).

## Methods

### Study design

We conducted a problem-based political economy analysis (PEA)(17,18), focusing on qualitative data from key informant interviews and a synthesis of published academic literature, grey literature and existing health system reports and policies. The PEA explored the intersection of regulatory policies, stakeholder influence, and economic factors that shape medical oxygen systems. This study formed one of the country case studies which informed the Lancet Global Health Commission on Medical Oxygen Security and had a specific focus on politics of medical oxygen system in the Peruvian setting.

### Setting

The country is highly centralized politically and economically in its capital city Lima, which accounts for around 1/3 of the total population, and 46% of the national gross domestic product (GDP) (19). Similarly, to other LA countries, Peru is considered as predominantly urban, with 20.7% of the population falling under the rural category (20). Nonetheless, lower respiratory infections remained the leading cause of death, followed by non-communicable diseases such as ischemic heart disease, chronic kidney disease, and diabetes, with chronic respiratory conditions accounting for 4.6% of deaths (21).

### Qualitative interviews

For the qualitative component, we identified and invited key informants who had actively participated in the design, financing, regulation, implementation, or advocacy of medical oxygen policies in Peru over the past five years. Participants were purposefully selected based on stakeholder mapping, with additional participants recruited through the snowball sampling method. Most interviews were conducted virtually via Zoom between June 7 and July 25, 2023. Three participants requested in-person interviews, all of which were conducted in a quiet space at a coffee shop. All invited key informants agreed to participate, and no compensation was provided. Interviews were conducted in Spanish by a social scientist experienced in in-depth interviewing, following a structured guide (Appendix 1). An assistant was also present to take notes. All interviews were recorded and transcribed.

### Document search

The published academic literature search was performed in the following databases: Medline, Embase, Web of Science and Global Index Medicus with the last search conducted on April 03, 2023, using the method described by Bramer et al (22). The search strategy was developed in Medline (Ovid) in collaboration with librarians at the Karolinska Institutet University Library. For each search concept Medical Subject Headings (MeSH-terms) and free text terms were identified. No language restriction was applied. The search was performed for several countries, we reviewed the papers related to Peru. Full search strategies for all databases are available in Appendix 2.

Gray literature was sourced using Google with three search terms: “Oxígeno medicinal Perú” (Medical oxygen Peru), “Oxígeno y COVID-19” (Oxygen and COVID-19), and “Planta de oxígeno medicinal Perú” (Medicinal oxygen plant Peru). The first 100 results for each term were reviewed, including documents and videos, while excluding advertisements, policy documents, and regulations.

A policy search was conducted using the Peruvian Ministry of Justice database (23) which provides open access to all national laws, norms, and regulations and features a search engine that allows filtering by date, type of norm, code, public sector, content, or keyword. We did not restrict the search by time frame or public sector, using the keyword “oxígeno medicinal” (medicinal oxygen).

### Analysis

The qualitative interviews were analyzed using the conceptual framework method (24). Two randomly chosen transcripts were critically read to identify initial themes and categories, considering the content of the text. Subsequently, the interviews were coded, with the aim of looking for similarities and differences in the discourses of the interviewees. For the document search we created a matrix for data abstraction. Each document was reviewed by two independent readers who generated short summaries of portions of the text which were introduced into the relevant cell of the matrix. The information from the different sources, qualitative interviews, literature and policies was integrated to create a narrative using the main themes found during the interviews.

### Ethics

The study protocol and instruments were reviewed and approved by the Cayetano Heredia University IRB (approval number 211379). Participants taking part in the key informant interviews provided informed consent before participation.

## Results

A total of 13 interviews were conducted including stakeholders from national or provincial governments, civil society, private sector, health providers, medical professional associations and multilateral agencies.

In the academic literature we found 631 articles, but only 7 were identified as relevant (see Prisma flow in Appendix 3). The grey literature review resulted in 87 pertinent documents (see Appendix 4) and in the policy review from 36 policies found, 23 were included in this analysis (Appendix 5).

### Peru contextual issues

Peru’s health system is highly fragmented, with distinct subsystems (public, social security (EsSalud) and private) operating independently under the oversight of the Ministry of Health (MINSA). Additionally, Peru is divided in 24 regions, each with its own government, budget and health directorate (decentralization). This segmentation has limited coordinated responses to public health needs, with medical oxygen as a critical example (25). One important contextual issue in Peru has been the political turmoil, in part associated to the corruption scandal in Latin America related to Odebrecht (26). As a result of the political instability, between 2016 to 2022, Peru has seen 7 different presidents (lasting on average just 8.5 months and the change of 17 Ministers of Health in that 6-year period, 8 of those during the pandemic years of 2020 and 2021 alone. Peru began its pandemic response preparations early and implemented several public health measures, aware of the weaknesses of its fragmented health system and the political situation (27). However, wide inequalities, with 70% of the population engaged in informal work with precarious income and social protection, poor housing and overcrowding, and an extended family culture in opposition to social distancing, all added to the perfect storm which resulted in very high rates of infection(28).

Despite MINSA’s role in regulating medical oxygen, the lack of a national oxygen strategy meant that each subsystem and each region operated its procurement and distribution channels, leading to inconsistencies and shortages during the COVID-19 crisis. The pandemic highlighted the limitations of the decentralized approach, as rural regions and areas outside Lima experienced severe oxygen shortages. Policies governing medical oxygen were not uniformly applied across regions, reflecting the underlying inequities exacerbated by fragmented governance.

### Regulatory and Economic Barriers to Oxygen Access: The policy trap

Although Peru approved the First National Policy for Medicines in 2004, the first essential medicinés list (EML) was not approved until 2010 (29). Before this, oxygen was classified as a medical device with no specific regulations. The 2010 EML introduced oxygen as a medication with a requirement of a concentration standard of >99% (30,31).

Starting in 2011, medical oxygen, like any other medicine, required a **sanitary registration** through DIGEMID (Peru’s national authority for medicines, medical devices, and diagnostics). Sanitary registration is the official process or documentation necessary for the legal approval and commercialization of pharmaceutical products in Peru. For medical oxygen, this process enforces standards for manufacturing, distribution, and labeling, including compliance with good manufacturing practices for oxygen production plants (32).

In 2012, INDECOPI (The Peruvian Institute for the Defense of Competition and the Protection of Intellectual Property) declared the >99% concentration requirement an illegal bureaucratic barrier, yet MINSA maintained this standard. The 2018 Ministerial Resolution updated the EML, still requiring >99% purity for medical oxygen. **Figure 1** summarizes the main regulations regarding medical oxygen in Peru, before the pandemic.

**Fig 1:**
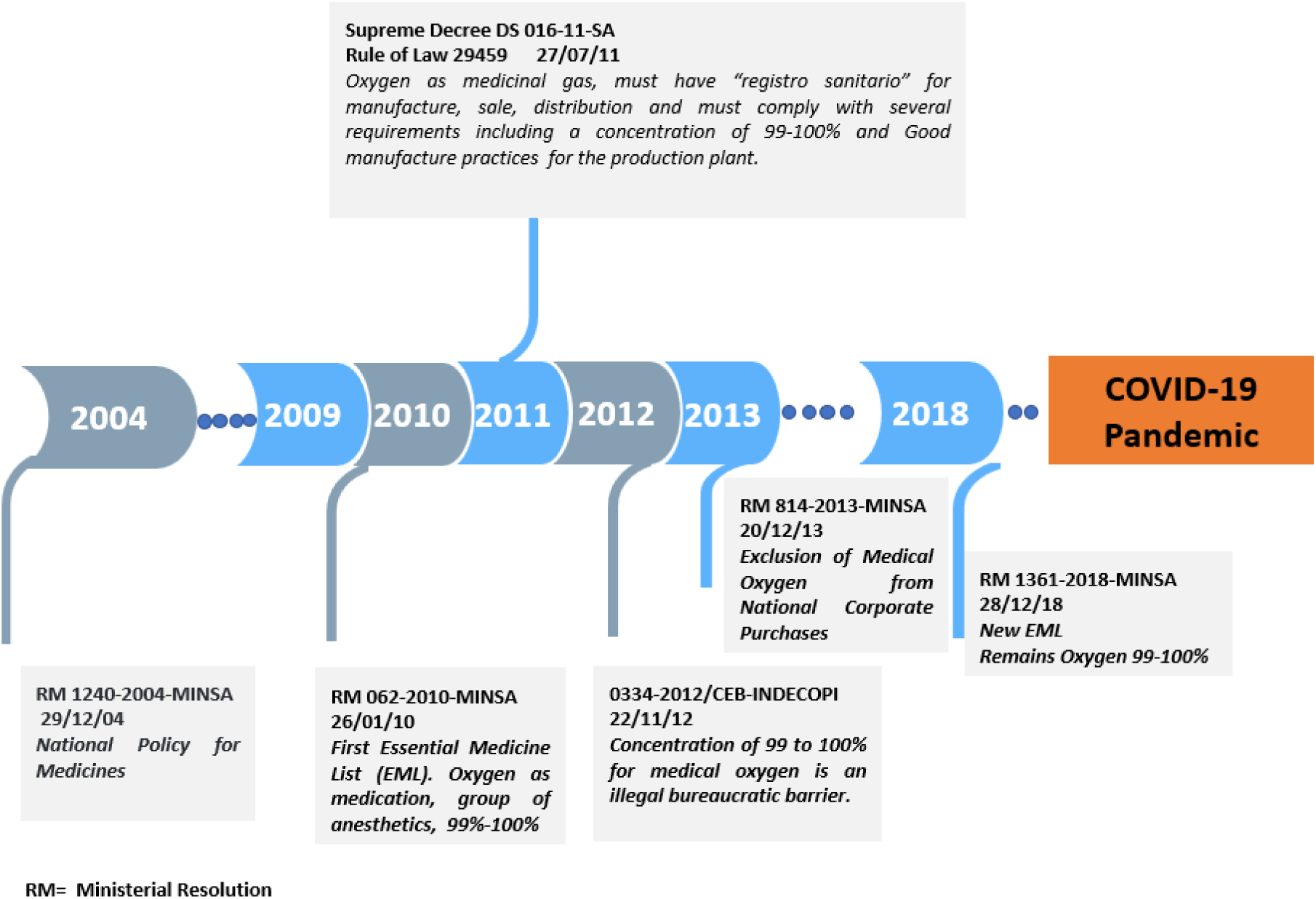
Timeline of the Regulations regarding Medical Oxygen in Peru before the COVID-19 Pandemic.

This led to six companies holding sanitary registrations to commercialize medical oxygen in Peru before the pandemic: Praxair Peru, Linde Gas Peru, Tecnogas, Air Products Peru, Indura Peru, and Oxyman Comercial. These companies are organized into two business groups, producing both industrial oxygen for the mining and metallurgical sectors (their primary clients) and medical oxygen. Together, these groups supplied 92.6% of the medical oxygen contracted by the government from 2008 to 2020, according to the State Contracting Supervisory Body (OSCE as per its Spanish acronym) (33–38). **(Table 1)**

**Table 1:** Main groups of Medical Oxygen in Peru contracting with the government: MINSA, EsSalud and regional governments, period 2008-2020*.

| Business group | Country | Peruvian company | Contracts (Million USD, %)** | Location of Production Plants *** | Type of oxygen |
| --- | --- | --- | --- | --- | --- |
| **Linde-Praxair**<br>**88.09 M USD**<br>**70.8%** | Germany | Praxair Peru | 56.23 (45.2%) | Callao | >99%<br>Production LOX |
|  |  | Linde Gas Peru | 20.93 (16.8%) | Ica |  |
|  |  | Tecnogas | 10.93 (8.8%) | Lima |  |
| **Air Products**<br>**27.17 M USD**<br>**21.8%** | USA | Air Products Peru | 17.80 (14.3%) | Chimbote | >99%<br>Production LOX |
|  |  | Indura Peru | 9.37 (7.5%) |  |  |
| Oxyman Comercial. | Peru | Oxyman Comercial | 4.06 (3.3%) | NA | >99%<br>Buys LOX and sells gaseous oxygen in cylinders |
| OxyCUSCO | Peru | OxyCUSCO | 0.20 (0.2%) | Cusco | >99% only in cylinders<br>Production |
| Oxigeno Iquitos | Peru | Oxigeno Iquitos | 1.0 (0.8%) | Loreto | 93% production |
| Oxigeno Loreto | Peru | Oxigeno Loreto | 1.59 (1.3%) | Ucayali | >99%, 93%****<br>Only sells gaseous oxygen in cylinders |
| Oxigeno Tao | Peru | Oxigeno Tao EIRL | 0.67 (0.5%) | Loreto | 93% production |
| Villalobos Rufino Ivan | Peru | Villalobos Rufino Ivan | 1.59 (1.3%) | San Martin | 93%<br>Sale and distribution of oxygen produce by "Oxigeno San Martin" |
| Total Million USD |  |  | 124.37 (100%) |  |  |

Source: Prepared with data for the three first business groups from reference (39) and from the Data base from the State Contracting Supervisory Body (OSCE). LOX=Liquid oxygen
\* We included only companies with registro sanitario before the pandemic.
\*\*Using exchange rate from 2008: 0.3124 USD per PEN
\*\*\* According to the DIGEMID Sanitary registration
\*\*\*\*Exception for regions with difficult access.

Interviewees also noted that pre-pandemic oxygen regulations in Peru were influenced by political and economic interests, requiring a >99% purity level despite the WHO’s recommendation of 93% or higher.

> ***“…(regarding the policy that changed the level of purity of medical oxygen)…it was a political and economic issue because, by raising the level of oxygen purity it was directed to two companies that were the only ones that did meet that requirement… it was like giving the way only to the two of them because the rest of the companies were not going to comply…”***
>
> ***National government***

Peru’s regulation requiring medical oxygen purity levels of >99% limited market competition by concentrating control within a few large suppliers, creating a “policy trap” that prevented smaller, more affordable providers from entering the market due to the costly technology needed to meet these standards.

Before the COVID-19 pandemic, according to interviewees, the country’s health establishments (HEs) had a sufficient supply of medical oxygen to meet patient demand. Due to varying conditions across HEs nationwide, oxygen distribution within facilities was managed in different ways: some used installed networks or purchased oxygen in cylinders, while others relied solely on rechargeable cylinders. Large HEs such as hospitals were responsible for procuring and replenishing their oxygen supplies. In smaller HEs, typically part of a health network, oxygen procurement was handled by the network administration or, in regional areas, by the regional government.

> ***“The supply of oxygen (before COVID) was sufficient, at least I have never been aware of a shortage that has put people’s lives at risk”***
>
> ***Health Provider***

> ***“… What we had at the time was enough for the patients who came to the hospital. At least in large hospitals, here in Lima, it seemed to me that we were in the right amount. In the periphery hospitals, in the regional hospitals, there is still no liquid medical oxygen tank (…) an ICU in a region or a province, works with oxygen cylinders and every four or six hours they have to be replaced, it became customary at that time and that’s how they have worked.”***
>
> ***Medical professional association***

### COVID-19 and Oxygen in Peru: limited market, slow response and lack of organization and data resulted in a crisis

In March 2020, Peru declared a mandatory quarantine, yet COVID-19 infections, hospitalizations, and deaths continued to rise. Oxygen became essential for critical COVID-19 patients, creating a major supply challenge for the government. The Ombudsman’s Office reported that at least 20% of hospitalized and ICU patients required oxygen, but supplies were insufficient (33). Initially, the full impact of COVID-19 as a respiratory illness and the demand for oxygen were underestimated, leading to a crisis as the health system became overwhelmed.

> ***“… In the months of June, July*** (***2020***)***, when the issue of oxygen was already complicated, we saw people in the streets with their cylinders trying to buy oxygen, and also, we saw people dying because of the lack of oxygen…”***
>
> ***Civil society***

Most high-purity oxygen plants are located along the central coast of Peru, while the few plants in the Andean and jungle regions produce only limited quantities of gaseous oxygen at 93% purity (38). High-purity packaging facilities are also primarily coastal, which led to major disparities in oxygen access. During the pandemic, these inequalities became clear, as Andean and jungle regions faced severe shortages—not only due to a lack of oxygen production but also due to limited distribution infrastructure to transport oxygen from coastal plants to these areas.

The main oxygen suppliers could not meet the rising oxygen demand in the country. In response, the government implemented policies to encourage local production and oxygen imports. For example, an emergency decree that temporarily authorized 93% oxygen purity and included measures to boost production and access. However, smaller companies struggled to obtain import permits or lacked the infrastructure to produce medical oxygen due to an absent market in prior years. The scarcity of 93% oxygen led to soaring prices and the emergence of an informal market, where oxygen cylinders were sold at exorbitant prices. Small legal and illegal vendors sold oxygen directly to the public, often using cylinders stolen from hospitals or cylinders not suitable for medical oxygen.

> ***“… I understand that the cost of a cubic meter of oxygen the real price is 8 soles, 8 to 10 soles and that at the time they sold the cylinder of 10 cubic meters up to 3000 soles, that is, the cubic meter was at 300 soles…”***
>
> ***Medical professional association***

However, officially lowering the oxygen purity standard to 93% required two laws, three ministerial resolutions, and seven months (**Figure 2**), highlighting bureaucratic delays potentially linked to economic and political factors as mentioned by the interviewees.

> ***“… Politics in general are based on interests (…) You didn’t even need a law declaring interest (law 31026), you didn’t even need a law that creates the National Oxygen Supply System (law 31113), because with the simple act of issuing a ministerial resolution, Ministers could have changed the requirement of oxygen purity from 99 to 93%. Why didn’t they do it? Isn’t it obvious? because they knew they were going to clash with some (economical) interests, and they preferred not to do it. Or they thought that the pandemic was already over, and they would not need to deal with the issue? Bah….”***
>
> ***National government***

**Fig 2:**
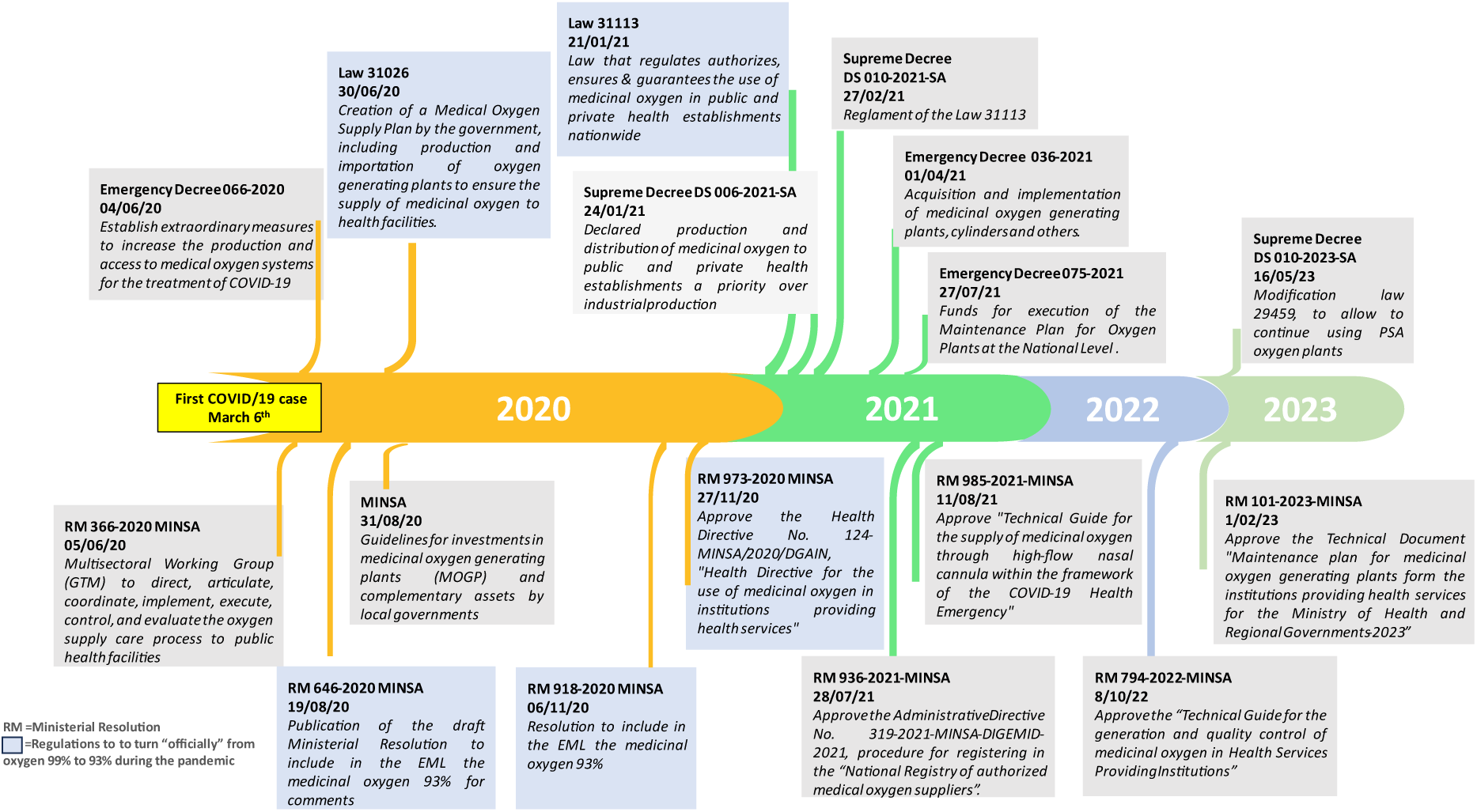
Timeline of the Regulations regarding Medical Oxygen in Peru during the COVID-19 Pandemic.

For the first time, national guidelines on medical oxygen use and quality control were issued one in 2021 and the other in 2022 (40,41).

Interviewees also suggested irregularities in oxygen procurement, both before and during the pandemic. Before COVID-19, hospitals purchased oxygen directly despite its designation as an essential medicine, which should have been centrally procured by the National Center for the Supply of Strategic Resources in Health from MINSA (CENARES) to reduce costs and improve access. In 2013, however, a ministerial resolution allowed health establishments to make fragmented, direct purchases, which increased costs and risked inefficiency and corruption (42).

Never in Peru before the pandemic, medical oxygen needs were addressed as a national problem that required a national solution or a national system. Interviewees from civil society and the business sector observed that authorities delayed in anticipating rising oxygen demand and ignored the need for a coordinated system. When oxygen shortages emerged, a Multisectoral Working Group (GTM) was formed to oversee oxygen supply nationwide (43), but interviewees noted that government response remained slow and ineffective, with insufficient production and a focus on intensive care unit (ICU) bed capacity rather than oxygen access.

> ***“… The ICU was full, more ICU beds were increasing, they needed more oxygen, that is, a problem was created, a big snowball (…) but the government insisted that we needed more ICU beds and nothing about the oxygen…”***
>
> ***Medical professional association***

It was evident that it was not possible to address the problem with the local production of oxygen. Eventually, $22 million was allocated to CENARES for oxygen procurement outside the country, but delays and bureaucratic obstacles led to the transfer of these responsibilities to other government agencies. Despite this shift, issues persisted, including problematic purchases such as oxygen cylinders that did not meet Peru’s national quality standards.

> ***“… Everything related to medical oxygen has been very opaque. While it is true that medical oxygen is in national demand (…) it should be bought by CENARES because it is considered a strategic medicine, however, it was bought by the hospitals themselves, due to a change in regulations. Then, when the pandemic came, “Peru Compras” bought it, “Legado” bought it, everyone bought the oxygen, but in the end, they bought and imported oxygen cylinders (…) the cylinders did not have the Peruvian quality standards, they did not have the Peruvian quality standard nanometers …***
>
> ***Civil Society***

Among public officials, many cited the absence of centralized leadership, lack of accountability, and high political turnover, which disrupted effective decision-making. Multiple government bodies were involved, but this fragmented structure hampered quick action and solutions. Some interviewees attributed this to personal interests and possible corruption.

Lack of data also worsened the oxygen crisis. Peru lacked accurate data on oxygen demand, production, and distribution, with no inventories of cylinders, isotanks, or oxygen plants in health facilities. Even during the pandemic, demand estimates were based only on case numbers and severity, limiting procurement efficiency.

> ***“… There was no national oxygen system or notification system, therefore, at the time of the pandemic, our capacity to respond to oxygen needs was not known… What we did was to walk blindly during the pandemic”***
>
> ***National government***

The 2021 Oxygen Law (31113) mandated that DIGEMID create a national registry for oxygen suppliers and stock, but this public registry, known as the “National Registry of Medical Oxygen – RENOXI Peru,” was not launched until September 2021, eight months later (44). Although the full database isn’t publicly accessible, most information can be found on the REUNIS platform (45), with some data available on a dedicated open-data site called “*oxígeno datos abiertos”*(46).

Interviewees raised concerns about the reliability of this data, pointing out that RENOXI lacks verification mechanisms to ensure data accuracy. In 2023, a cyberattack further compromised RENOXI’s registry, resulting in data loss and further diminishing confidence in the system.

> ***“…I don’t know if what RENOXI reports to me (data) is really true… there the consumption is based on the number of cylinders which are delivered from the pharmacy service, if I deliver 3 cylinders of 10 m³ it means that 30 m³ have been consumed, but It is not that they have been consumed, they are just being delivered, consumption is probably lower… or they may forget to register, and nobody supervises or rechecks the numbers”***
>
> ***Regional government***

> ***“… At the beginning of 2023, the website (RENOXI) went down, due to a computer attack, and the information was lost…”***
>
> ***National government***

### Oxygen plants, civil society and private sector contributions

In August 2020, MINSA issued guidelines for local governments on investing in oxygen plants and equipment, allocating funds for the purchase and installation of oxygen plants and cylinders (47). However, government procurement was slow. Civil society, including academia, NGOs, the private sector outside the oxygen industry, and notably the Catholic Church (with some regional interfaith coalitions), played a crucial role in addressing the government’s inability to procure and implement oxygen plants. Through fundraising efforts, these groups donated oxygen plants, cylinders, and oxygen supplies, significantly improving access in areas where government support was insufficient.

Nationally, *Respira Perú*, an initiative led by the Peruvian Episcopal Conference, the National Society of Industries and other parties, was the most prominent civil society effort, raising funds to purchase and install oxygen supplies and plants. In the regions, local groups organized similar initiatives as COVID-19 cases surged, sometimes with support from political figures and private companies. Many plants obtained through these efforts were donated to hospitals and health establishments managed by MINSA, regional governments, or municipalities. For Respira Perú, a medical council—including representatives from MINSA, EsSalud, and leading doctors—helped decide where to install plants based on need.

However, not all the initiatives were successful. Some interviewees noted that DIGEMID’s bureaucratic demands and limited staff hindered plant approvals, leaving oxygen plants stalled at customs without authorization.

> ***“…there was a lot of informality during the pandemic… many oxygen generating plants were set up and began to operate without having any type of authorization or anything, because people were simply dying…”***
>
> ***Multilateral agency***

> ***“…they [the government officials] put the noose around their necks because they began to demand a requirement, and they did not have officials to carry out the verifications. They issued the documents at the wrong time… we had the oxygen plants at customs, and we could not remove them because there was no authorization from DIGEMID…”***
>
> ***Civil Society***

Despite successful oxygen plant installations, operational challenges arose due to high staff turnover, limited training and lack of replacement parts and maintenance plans. By the end of the pandemic, 496 oxygen plants had been installed across the country, funded by national, regional, and private donations.

### The situation of medical oxygen after COVID-19 and lack of preparedness for future pandemics

The process of donating, distributing, and implementing oxygen plants in Peru was marked by opportunistic actions without plans for maintenance or replacement. The allocation was not strategically directed to high-need areas, leading to inequitable distribution, and many plants were left in legal limbo, preventing government investment in their upkeep.

As a result, nearly 200 oxygen plants nationwide are now unused, broken, or abandoned. Since oxygen demand declined post-pandemic, the operation of these plants now depends on local health establishment requirements.

According to one interviewee, MINSA has allocated funds for oxygen plant maintenance since 2021, including for both purchased and donated plants. Yet only part of this budget has been utilized due to delays in legally clearing these assets, which should typically take no more than 90 days.

> ***“Up to 3 maintenance plans were made for the oxygen plants: in 2021, 2022 and 2023, and 22 million soles were programmed for it… But I don’t think that money has been executed. There are a lot of problems. For example, at the Loayza Hospital there are 3 oxygen plants that were donated, one by the Southern Mining company. The problem is that you can’t do anything, you can’t invest government funds for maintenance because they are not in someone’s name, nothing is formalized. That’s a knot…”***
>
> ***National government***

Changes in authorities and varying levels of interest or awareness have delayed the legal clearance process. Additionally, some believe that maintenance delays might be linked to potential corruption in medical oxygen procurement. Regional barriers include staff lacking proper training in plant maintenance and a shortage of service providers. Consequently, health establishments are mostly reverting to pre-pandemic practices, purchasing oxygen from a few primary suppliers instead of maintaining their plants.

> ***“… in Peru, for reasons that need to be investigated, because the truth is that I have heard different versions of the origin, the individual cylinder is used a lot, which is more expensive, more inefficient… but it also lends itself much more easily to dishonest practices (…) I don’t know, but what I can say is that today I know that there are hospitals that are ceasing to use oxygen plants to return to the oxygen cylinder system… and they’re letting the plant go bad…”***
>
> ***Private Sector***

Other barriers, more common in the regions, include staff lacking proper training for plant maintenance and a shortage of service providers. Consequently, health establishments are mostly reverting to pre-pandemic practices, purchasing oxygen from a few primary suppliers instead of maintaining their plants. In the 2024 Public Sector Budget Law (Law 31953), about USD 5.7 million has been earmarked for preventive and corrective maintenance for medical oxygen plants, though no clear maintenance plan is in place (48).

When asked about preparedness for future pandemics, most interviewees expressed that despite measures taken during COVID-19—such as providing oxygen plants and concentrators—Peru has not effectively learned from the experience. They believe similar issues would arise in future health crises.

> ***“… If we were to face an epidemic again, that kind of circumstances would be very similar, we have not learned the lesson, maybe a little bit or to some degree a better response, but we would still collapse in our need for oxygen.”***
>
> ***Health Provider***

At a management level, interviewees emphasized the need for a National Oxygen System to prepare for emergencies, including robust demand-supply tracking, forecasting, equipment inventories, maintenance, operation supervision, and quality control, with equitable oxygen distribution.

> ***“… The Executive issued (finally!) an Emergency Decree 066, which says that oxygen drops from 99% to 93% (concentration), that was good. But I want to warn you that this is not the essential issue… The essential thing is to create a National oxygen system, which could strategically assess the demand, assure the supply, create an oxygen quality control system, and understand and assure the equipment, infrastructure, networks, distribution, training etc. etc (…) if we don’t have that, it’s not going to work…”***
>
> ***National government***

Several interviewees support maintaining operational oxygen plants at strategic locations across the country, considering it economically advantageous in the long term. However, some also expressed concerns about the operational and maintenance demands relative to the potential benefits, highlighting the need for further economic studies. All interviewees emphasized the importance of securing local oxygen availability to reduce dependency on external suppliers.

> ***“…strategic oxygen plants would surely be a saving for the Government. Nobody has done the math. If you invest in maintenance and operational plants, and adequate well-trained personnel, that is surely a saving and a good investment for the country…”***
>
> ***National government***

Addressing future pandemics will also require a stronger focus on human resources. Participants highlighted the need for trained technical personnel to ensure continuous oxygen supply, as well as for dedicated professionals capable of managing critical situations, and improve the training of health professionals on rational oxygen use. They stressed that retaining skilled staff across administrative changes is essential for continuity in crisis response processes.

> ***“ We need medical oxygen in all the country, we need good distribution systems, networks, trained providers and technical personnel. People die due to the lack of oxygen, but we are not counting.”***
>
> ***National government***

> ***“… I think that in any case it is necessary to have more technical staff for maintenance of the plant and support, unfortunately the hospital does not have the capacity to be able to hire more people and there are very few people with a good level of qualification for those positions, but it would require personnel because there is a need to have teams that work 24 hours a day and in great demand so you have to have a whole technical support and maintenance staff…”***
>
> ***Health Provider***

## Discussion

This study reveals significant barriers to equitable oxygen access in Peru, reflecting broader challenges faced by low- and middle-income countries. The COVID-19 pandemic exposed Peru’s lack of preparedness, stemming from longstanding regulatory and logistical weaknesses. While medical oxygen was designated an essential medicine, the <>99% purity requirement created a “policy trap,” limiting competition and concentrating market control within a few large suppliers. The WHO International Pharmacopoeia defines medicinal oxygen as Oxygen 99.5% or Oxygen 93% that contains not less than 90.0% and not more than 96.0% (v/v) of O_2_, or other products with different oxygen concentrations and/or produced using different production methods, if approved by the appropriate national or regional authority (49). This regulatory barrier prevented smaller, potentially more affordable providers from entering the market, contributing to severe shortages during the pandemic.

The fragmented and decentralized health system in Peru further complicated oxygen distribution, particularly in rural regions. While the Ministry of Health oversees medical oxygen policies, the lack of a national oxygen strategy and centralized procurement framework led to inconsistencies in supply and an inefficient crisis response. Government entities responded slowly, hindered by bureaucratic delays and limited coordination. Even as COVID-19 cases surged, the focus remained on ICU bed capacity rather than securing sufficient oxygen supplies, highlighting a misalignment in priorities during critical moments.

The experience in Peru during the COVID-19 pandemic highlights a broader issue regarding equipment donations in emergency responses. In Peru, civil society and the private sector played pivotal roles in addressing oxygen shortages by donating oxygen plants and essential supplies. However, the lack of a structured maintenance plan resulted in many of these donated plants falling into disrepair or disuse. This scenario underscores a critical flaw in the global approach to medical equipment donations: while initial donations can provide immediate relief, their long-term effectiveness depends on robust systems for maintenance, training, and integration into local health infrastructure. Without these systems, the impact of such donations diminishes, leaving communities vulnerable when the equipment inevitably degrades or breaks down.

This issue extends beyond Peru, as evidenced during the COVID-19 pandemic when initiatives like the Access to COVID-19 Tools Accelerator (ACT-A) raised over $1 billion, much of which was spent on equipment donations. However, as noted in Unitaid’s Global Oxygen Strategic Framework, sustainable planning and investment in medical oxygen systems require far more than initial donations; an estimated $4 billion is needed to develop comprehensive and durable oxygen delivery systems in low- and middle-income countries(50). Similarly, research published emphasizes the dangers of poorly planned or inappropriate donations, which can overwhelm recipient health systems, lead to waste, or create dependency without ensuring long-term benefits. These examples highlight the need for a paradigm shift in how donations are approached, moving from short-term crisis responses to investments in sustainable systems that empower local health care infrastructure to manage and maintain critical resources independently (51–53).

Our findings underscore the necessity for Peru to establish a National Oxygen System to address the recurring challenges of oxygen shortages and the sustainability of donated equipment. Such a system should integrate supply chain management, maintenance funding, and infrastructure investments to ensure a reliable, locally available oxygen supply. A comprehensive approach would also require training personnel to operate and maintain these systems, reducing dependency on external suppliers and enhancing resilience against future health crises. This proactive strategy would not only address immediate needs but also strengthen Peru’s health system capacity to withstand future pandemics or emergencies that demand oxygen availability.

This recommendation aligns with the World Health Organization’s recent resolution on strengthening health systems for pandemic preparedness, as highlighted in the document adopted during the 152nd Executive Board Meeting (54). The resolution emphasizes the need for countries to take strategic actions to ensure access to critical medical resources, including oxygen, as part of broader efforts to achieve universal health coverage and build resilient health systems. It explicitly calls for investments in supply chains, infrastructure, and human resources to address gaps in essential medical supplies, particularly in low- and middle-income countries. Peru’s experience during the COVID-19 pandemic serves as a clear example of why these measures are vital, as the absence of a structured system for oxygen supply left many donated plants in disrepair and communities vulnerable.

By establishing a National Oxygen System, Peru would not only meet the immediate recommendations outlined in the WHO resolution but also set a precedent for other countries facing similar challenges. This system would ensure that oxygen resources are not only available but also sustainable in the long term, reducing reliance on ad hoc donations and external support. Such an initiative would represent a significant step forward in achieving equitable access to essential medical supplies and safeguarding public health in the face of future global health crises.

In conclusion, addressing the regulatory, logistical, and operational gaps in Peru’s oxygen supply system is critical. Developing a coordinated, well-regulated national oxygen framework could prevent similar crises in the future and safeguard public health during emergencies.

## Data Availability

The datasets used and/or analyzed during the current study are available from the corresponding author upon reasonable request.

## Ethical approval and consent to participate

The research was reviewed and approved by the Cayetano Heredia University IRB (approval number 211379). Written informed consent to participate in the study was obtained from all human research participants.

## Authors’ contributions

PJG, FEK, FS and CK designed and conceptualized the study. PJG, FEK and CK developed the study protocol. PJG, JMG and LN did the data collection, transcription, data management and preliminary analysis of the data. PJG, FEK, JMG, LN and CK contributed to the data analysis, and report writing. All authors contributed to the interpretation of the findings. PJG wrote the first draft of the paper. FEK, JMG, LN, FS and CK reviewed, revised, and contributed to writing to the paper. All authors read and approved of the final manuscript. PJG, FEK, JMG, LN, FS and CK read and met the ICMJE criteria for authorship.

## Acknowledgments

The authors express their gratitude to the Lancet Global Health Commission on Medical Oxygen Security, Commissioners and Advisors of work package 4 – financing and political economy. Additionally, the authors acknowledge the research librarians at the Karolinska Institute, who conducted literature searches and retrieved articles for this case study as part of the Lancet Global Health Commission on Medical Oxygen Security.

