## Appendix4 for "Challenges in the medical oxygen ecosystem of Peru: a political economy analysis"

### Slide 1
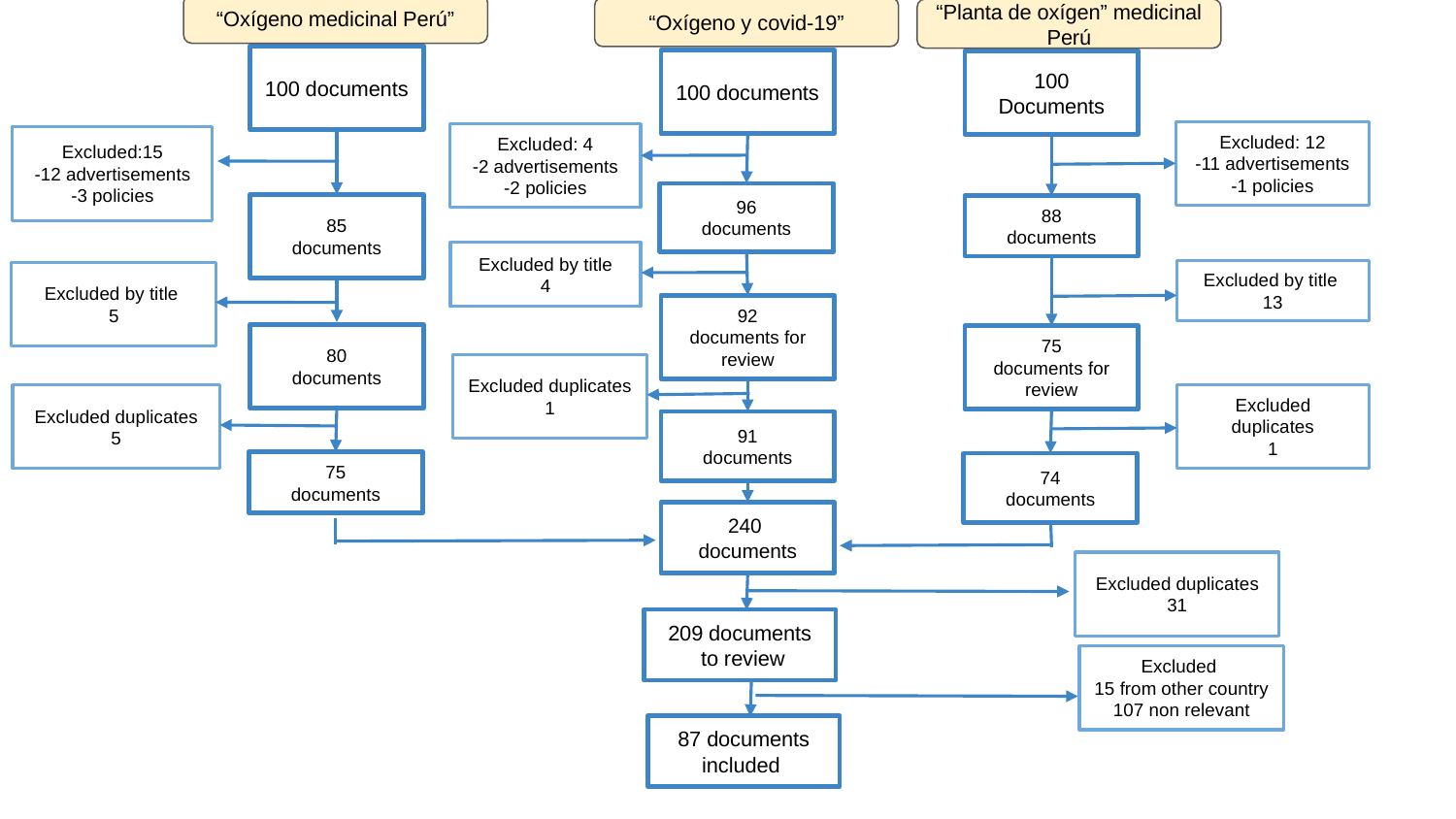

“Oxígeno medicinal Perú”
“Oxígeno y covid-19”
“Planta de oxígen” medicinal Perú
100 documents
100 documents
100 Documents
Excluded: 12
-11 advertisements
-1 policies
Excluded: 4
-2 advertisements
-2 policies
Excluded:15
-12 advertisements
-3 policies
96
documents
85
documents
88
documents
Excluded by title
4
Excluded by title
13
Excluded by title
5
92
documents for review
80
documents
75
documents for review
Excluded duplicates
1
Excluded duplicates
1
Excluded duplicates
5
91
documents
75
documents
74
documents
240
documents
Excluded duplicates
31
209 documents
 to review
Excluded
15 from other country
107 non relevant
87 documents included
