## Appendix5 for "Challenges in the medical oxygen ecosystem of Peru: a political economy analysis"

### Slide 1
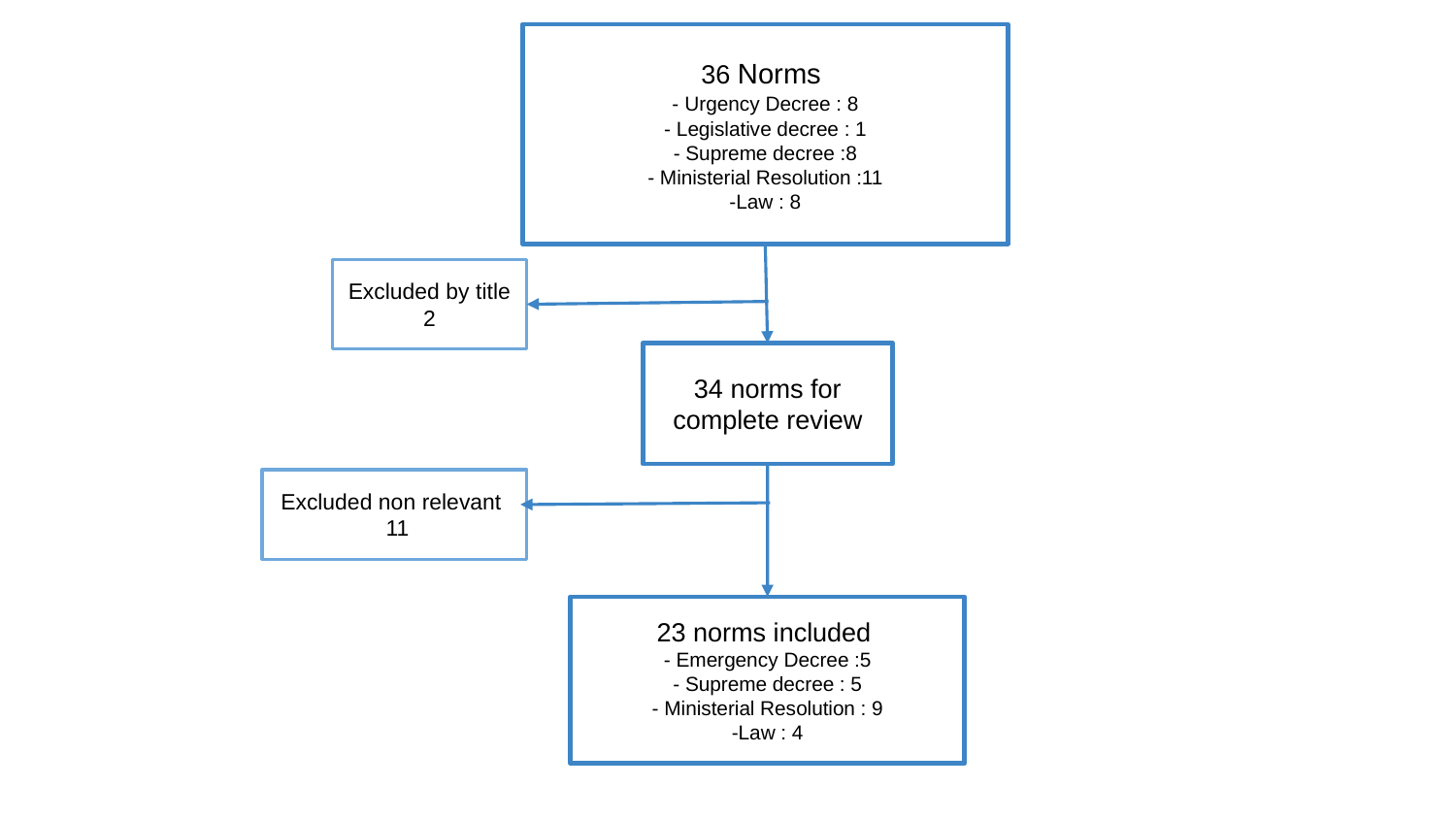

36 Norms
- Urgency Decree : 8
- Legislative decree : 1
- Supreme decree :8
- Ministerial Resolution :11
-Law : 8
Excluded by title
2
34 norms for complete review
Excluded non relevant
 11
23 norms included
- Emergency Decree :5
- Supreme decree : 5
- Ministerial Resolution : 9
-Law : 4
