## Appendix1 for "Challenges in the medical oxygen ecosystem of Peru: a political economy analysis"

**Appendix 1**

**Interview Guide**

**Introduction and Consent**

Introduce yourself, confirm that the participant has received the study’s written information, and verbally explain the study’s objectives, confidentiality, and their right to withdraw. Allow time for any clarifications before obtaining informed consent. If they agree to continue with the interview, inform them that recording will begin.

**Participant Experience and Role**

- Tell me a bit about your role in medical oxygen services.
- How long have you been in this position?
- Has this role changed in recent years (e.g., in relation to COVID-19)?

**Oxygen Policies**

- Can you discuss the relevant policies you work with around medical oxygen services?

*Probe: Who published this policy? Is it institutional, national, regional, or global? Was there a political, scientific, economic, or logistical reason for it?*

- If no policy is provided, is this due to nonexistence or irrelevance to your work?
- Are these guidelines/policies reviewed or evaluated?
- How frequently? How? By whom?
- Do you think the current medical oxygen policies are sufficient?
- Why or why not?

**Financing: Reflecting on Funding for Medical Oxygen Services**

- Where does funding for medical oxygen come from?
- Who are the main actors responsible for funding oxygen programs?
- Are they different from regional-level actors?
- How is healthcare funding prioritized?
- What challenges exist in securing national or local government commitment to oxygen programs?
- What challenges exist in obtaining private (for-profit or nonprofit) funding for local oxygen programs?
- What is the main shortfall in resources for oxygen programs?
- Infrastructure, consumables, personnel?

**Regulation, Accountability, and Monitoring: Oversight Mechanisms for Medical Oxygen Services**

- What accountability mechanisms exist for medical oxygen safety within the healthcare system?
- Who are the main actors responsible for implementing these?
- Who is responsible for regulating medical oxygen?
- Includes clinical delivery, production, diagnostic devices.
- Are these actors different from regional ones?
- Can you describe the monitoring mechanisms?
- Do you face any regulatory challenges?

**Policy Commitment**

- In your view, what are the key areas any new oxygen policy should focus on? For example, technological approaches, healthcare system strengthening.
- Which areas do you think the current policy addresses well (i.e., areas that do not need change)?
- Which areas require further development? Why? How could this be done?

**Additional questions**

- What challenges were faced regarding oxygen during the pandemic?
- Can you explain how or why these issues occur?
- Who were the key actors? What actions were taken?
- What was done across sectors to address these challenges? Was it successful or not? What obstacles were there? How were they overcome (or not)?
- What is the situation today? What has been learned and what has changed?
- In your opinion, if a pandemic occurred now, what would the oxygen situation be?
- What key lessons could you share?
