## Appendix2 for "Challenges in the medical oxygen ecosystem of Peru: a political economy analysis"

1. Medline

| Interface: Ovid MEDLINE(R) and Epub Ahead of Print, In-Process & Other Non-Indexed Citations and Daily  Date of Search: 31 March 2023  Number of hits: 3620  Comment: In Ovid, two or more words are automatically searched as phrases; i.e. no quotation marks are needed | Field labels   - exp/ = exploded MeSH term - / = non exploded MeSH term - .ti,ab,kf. = title, abstract and author keywords - adjx = within x words, regardless of order - * = truncation of word for alternate endings |
| --- | --- |
| Ovid MEDLINE(R) ALL <1946 to March 30, 2023> 1 \| exp Oxygen Inhalation Therapy/ \| 28246; 2 \| exp Oximetry/ \| 16591; 3 \| Oxygen/sd \| 107; 4 \| Oxygen/st \| 32; 5 \| Oxygen/tu \| 3680; 6 \| oxygen*.ti,ab,kf. \| 671667; 7 \| oximet*.ti,ab,kf. \| 16396; 8 \| Medical oxygen.ti,ab,kf. \| 116; 9 \| or/1-8 \| 691298; 10 \| COVID-19/ or exp COVID-19 Testing/ or COVID-19 Vaccines/ or SARS-CoV-2/ \| 222928; 11 \| (coronavirus/ or betacoronavirus/ or coronavirus infections/) and (disease outbreaks/ or epidemics/ or pandemics/) \| 40224; 12 \| (nCoV* or 2019nCoV or 19nCoV or COVID19* or COVID or SARS-COV-2 or SARSCOV-2 or SARS-COV2 or SARSCOV2 or SARS coronavirus 2 or Severe Acute Respiratory Syndrome Coronavirus 2 or Severe Acute Respiratory Syndrome Corona Virus 2).ti,ab,kf. \| 335614; 13 \| (longCOVID* or postCOVID* or postcoronavirus* or postSARS*).ti,ab,kf. \| 81; 14 \| ((coronavirus* or corona virus* or betacoronavirus*) adj3 (pandemic* or epidemic* or outbreak* or crisis)).ti,ab,kf. \| 15253; 15 \| or/10-14 \| 347854; 16 \| 9 or 15 \| 1029755; 17 \| Sweden/ \| 81195; 18 \| (sweden* or swedish).ti,ab,kf. \| 84949; 19 \| Bangladesh/ \| 14154; 20 \| bangladesh*.ti,ab,kf. \| 20510; 21 \| Malawi/ \| 6627; 22 \| (Malawi* or Nyasaland).ti,ab,kf. \| 9360; 23 \| Nigeria/ \| 33632; 24 \| Nigeria*.ti,ab,kf. \| 42246; 25 \| \| 10654; 26 \| (peru or peruv*).ti,ab,kf. \| 17061; 27 \| exp India/ \| 117961; 28 \| india*.ti,ab,kf. \| 202744; 29 \| or/17-28 \| 448196; 30 \| exp "Health Care Economics and Organizations"/ \| 1677889; 31 \| exp Guidelines as Topic/ \| 172708; 32 \| exp Legislation as Topic/ \| 174737; 33 \| exp Policy/ \| 191489; 34 \| Decision Making/ \| 104139; 35 \| Decision Making, Shared/ \| 1853; 36 \| Decision Making, Organizational/ \| 11238; 37 \| exp Industry/ \| 349838; 38 \| exp Health Planning/ or Delivery of Health Care/ \| 463497; 39 \| (policy* or policies).ti,ab,kf. \| 361907; 40 \| (legislat* or law*).ti,ab,kf. \| 200096; 41 \| (social control or regulation* or guideline*).ti,ab,kf. \| 1493264; 42 \| decision making.ti,ab,kf. \| 189918; 43 \| ((drug or health or healthcare or health care or medical) adj2 (sector* or industr* or market* or financ* or planning* or program* or reform*)).ti,ab,kf. \| 142218; 44 \| or/30-43 \| 3980854; 45 \| 16 and 29 and 44 \| 3620 | |

2. Embase

| Interface: embase.com  Date of Search: 3 April 2023  Number of hits: 3953  Comment: Emtree is the controlled vocabulary in Embase | Field labels   - /exp = exploded Emtree term - /de = non exploded Emtree term - ti,ab,kw = title, abstract and author keywords - NEAR/x = within x words, regardless of order - * = truncation of word for alternate endings |
| --- | --- |
| No. Query Results #41 \| #13 AND #27 AND #40 \| 3953; #40 \| #28 OR #29 OR #30 OR #31 OR #32 OR #33 OR #34 OR #35 OR #36 OR #37 OR #38 OR #39 \| 561915; #39 \| india*:ti,ab,kw \| 277211; #38 \| 'india'/exp OR 'indian'/de \| 219130; #37 \| peru:ti,ab,kw OR peruv*:ti,ab,kw \| 21910; #36 \| 'peru'/de OR 'peruvian'/de \| 16326; #35 \| nigeria*:ti,ab,kw \| 52676; #34 \| 'nigeria'/de OR 'nigerian'/de \| 47503; #33 \| malawi*:ti,ab,kw OR nyasaland:ti,ab,kw \| 10935; #32 \| 'malawi'/de OR 'malawian'/de \| 9772; #31 \| bangladesh*:ti,ab,kw \| 24417; #30 \| 'bangladesh'/de OR 'bangladeshi'/de \| 21929; #29 \| sweden*:ti,ab,kw OR swedish:ti,ab,kw \| 110823; #28 \| ('sweden'/de OR swedish) AND 'citizen'/de \| 0; #27 \| #14 OR #15 OR #16 OR #17 OR #18 OR #19 OR #20 OR #21 OR #22 OR #23 OR #24 OR #25 OR #26 \| 4226178; #26 \| ((drug OR health OR healthcare OR 'health care' OR medical) NEAR/2 (sector* OR industr* OR market* OR financ* OR planning* OR program* OR reform*)):ti,ab,kw \| 173477; #25 \| 'decision making':ti,ab,kw \| 259190; #24 \| 'social control':ti,ab,kw OR regulation*:ti,ab,kw OR guideline*:ti,ab,kw \| 1988646; #23 \| legislat*:ti,ab,kw OR law*:ti,ab,kw \| 238114; #22 \| policy*:ti,ab,kw OR policies:ti,ab,kw \| 424491; #21 \| 'health care delivery'/de \| 206325; #20 \| 'health care planning'/exp \| 112231; #19 \| 'industry'/exp \| 365675; #18 \| 'decision making'/exp \| 455296; #17 \| 'policy'/exp \| 325726; #16 \| 'law'/exp \| 123509; #15 \| 'practice guideline'/de \| 533695; #14 \| 'health care cost'/exp \| 334863; #13 \| #6 OR #12 \| 785625; #12 \| #/ OR #8 OR #9 OR #10 OR #11 \| 402215; #11 \| ((coronavirus* OR 'corona virus*' OR betacoronavirus*) NEAR/3 (pandemic* OR epidemic* OR outbreak* OR crisis)):ti,ab,kw \| 15164; #10 \| longcovid*:ti,ab,kw OR postcovid*:ti,ab,kw OR postcoronavirus*:ti,ab,kw OR postsars*:ti,ab,kw \| 7747; #9 \| ncov*:ti,ab,kw OR 2019ncov:ti,ab,kw OR 19ncov:ti,ab,kw OR covid19*:ti,ab,kw OR covid:ti,ab,kw OR 'sars cov 2':ti,ab,kw OR 'sarscov 2':ti,ab,kw OR 'sars cov2':ti,ab,kw OR sarscov2:ti,ab,kw OR 'sars coronavirus 2':ti,ab,kw OR 'severe acute respiratory syndrome coronavirus 2':ti,ab,kw OR 'severe acute respiratory syndrome corona virus 2':ti,ab,kw \| 369309; #8 \| ('coronavirinae'/de OR 'betacoronavirus'/de OR 'coronavirus infection'/de) AND ('epidemic'/de OR 'pandemic'/de) \| 11515; #7 \| 'coronavirus disease 2019'/de OR 'covid-19 testing'/exp OR 'sars-cov-2 vaccine'/de OR 'severe acute respiratory syndrome coronavirus 2'/de \| 322811; #6 \| #1 OR #2 OR #3 OR #4 OR #5 \| 400226; #5 \| oximet*:ti,ab,kw \| 23791; #4 \| oxygen*:ti,ab,kw \| 821258; #3 \| 'oxygen'/de \| 257430; #2 \| 'oximetry'/exp \| 35192; #1 \| 'oxygen therapy'/exp \| 104359 | |

4. Web of Science Core Collection

| Interface: Clarivate Analytics  Editions = A&HCI , ESCI , SCI-EXPANDED , SSCI  Date of Search: 3 April 2023  Number of hits: 4290 | Field labels   - TS/Topic = title, abstract, author keywords and Keywords Plus - NEAR/x = within x words, regardless of order - * = truncation of word for alternate endings   Note: the *Exact search*-function was used for all the searches |
| --- | --- |
| # Search Query Results 1 \| "TS=(oxygen*)" \| 1360186; 2 \| "TS=oximet*" \| 17384; 3 \| "#1 OR #2" \| 1367747; 4 \| "TS=(nCoV* OR 2019nCoV OR 19nCoV OR COVID19* OR COVID OR SARS-COV-2 OR SARSCOV-2 OR SARS-COV2 OR SARSCOV2 OR ""SARS coronavirus 2"" OR ""Severe Acute Respiratory Syndrome Coronavirus 2"" OR ""Severe Acute Respiratory Syndrome Corona Virus 2"" )" \| 410468; 5 \| "TS=(longCOVID* OR postCOVID* OR postcoronavirus* OR postSARS* )" \| 187; 6 \| "TS=((coronavirus* OR ""corona virus*"" OR betacoronavirus* ) NEAR/2 (pandemic* OR epidemic* OR outbreak* OR crisis ))" \| 19278; 7 \| "#4 OR #5 OR #6" \| 414464; 8 \| "#3 OR #7" \| 1773100; 9 \| "TS=(policy* OR policies )" \| 1104049; 10 \| "TS=(legislat* OR law* )" \| 872196; 11 \| "TS=(""social control"" OR regulation* OR guideline* )" \| 2070976; 12 \| "TS=""decision making""" \| 414003; 13 \| "TS=((drug OR health OR healthcare OR ""health care"" OR medical ) NEAR/1 (sector* OR industr* OR market* OR financ* OR planning* OR program* OR reform* ))" \| 131861; 14 \| "#9 OR #10 OR #11 OR #12 OR #13" \| 4298656; 15 \| "TS=(sweden* OR swedish )" \| 152988; 16 \| "TS=bangladesh*" \| 41142; 17 \| "TS=(Malawi* OR Nyasaland )" \| 16742; 18 \| "TS=Nigeria*" \| 70492; 19 \| "TS=(peru OR peruv* )" \| 47350; 20 \| "TS=india*" \| 503100; 21 \| "#15 OR #16 OR #17 OR #18 OR #19 OR #20" \| 819351; 22 \| "#8 AND #14 AND #21" \| 4290 | |

5. WHO Global Index Medicus

| Interface: Global Index Medicus web interface https://www.globalindexmedicus.net/  Date of Search: 3 April 2023  Number of hits: 667 | Field labels   - Tw = Title, Abstract, Subject |
| --- | --- |
| (Oxygen OR oximet* OR "covid 19" OR corona*) AND (Policy OR Policies OR Regulaton* OR guideline* OR Legislat* OR law* OR reform* OR finance* OR industr* OR market* OR sector* Or planning OR program*) AND (Sweden* OR Swedish OR Bangladesh* OR Malawi* OR Nyasaland OR Nigeria* OR Peru OR Peruv* OR india*) | |
