## Appendix3 for "Challenges in the medical oxygen ecosystem of Peru: a political economy analysis"

### Slide 1
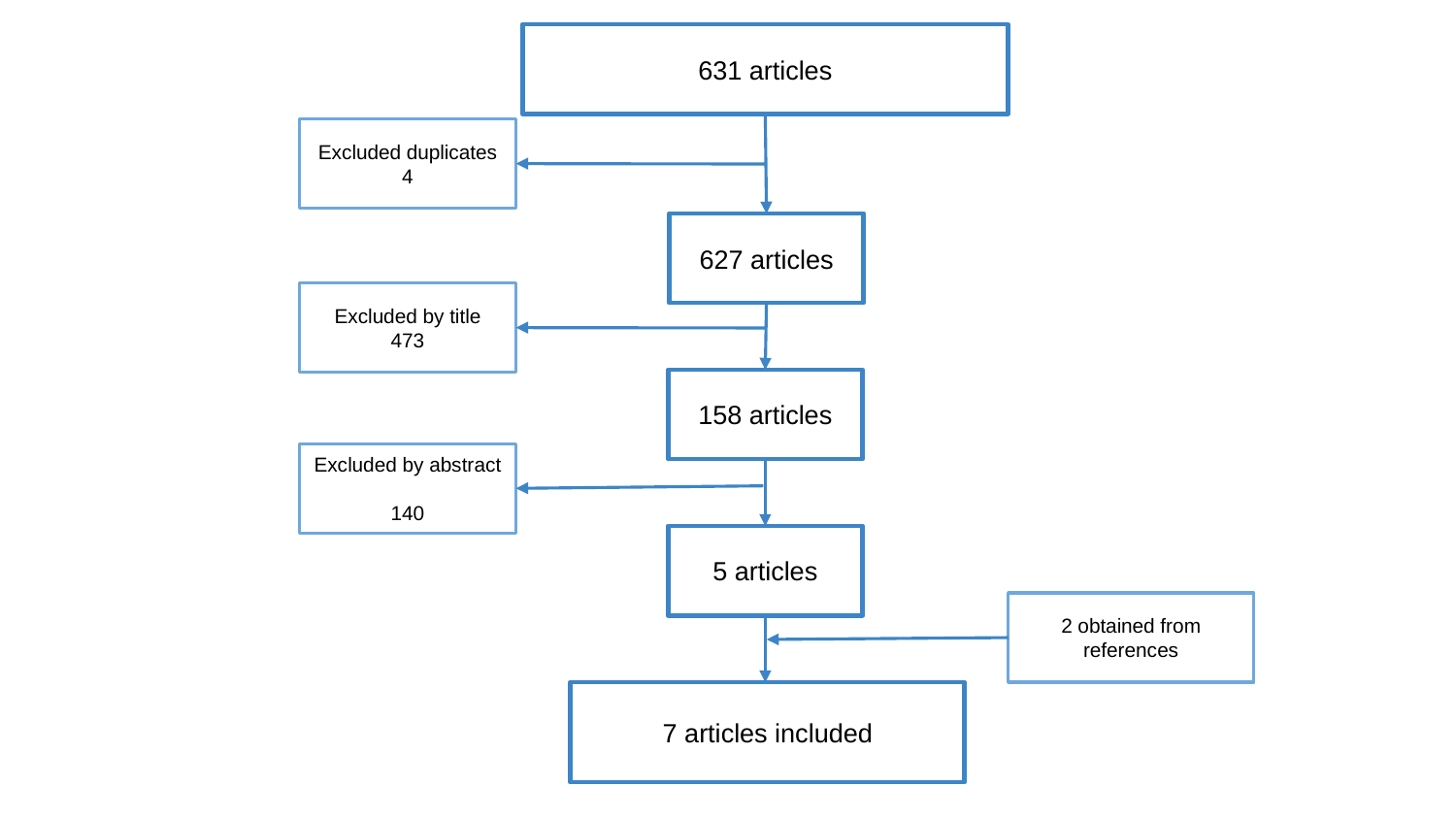

631 articles
Excluded duplicates
4
627 articles
Excluded by title
473
158 articles
Excluded by abstract
140
5 articles
2 obtained from references
7 articles included
